# Predictors of Supplemental Opioid Use After Third Molar Extraction

**DOI:** 10.1101/2025.07.18.25331797

**Authors:** Neeraj Panchal, Steven Wang, Rania A. Habib, Brian P. Ford, Truongan Pham, Perla M. Cherfane, Gaurav Gupta, Sydney A. Juska, Stacey A. Secreto, Carsten Skarke, Gregory R. Grant, Elliot V. Hersh, John T. Farrar, Tilo Grosser, Katherine N. Theken

**Affiliations:** Department of Oral and Maxillofacial Surgery and Pharmacology, School of Dental Medicine, University of Pennsylvania, Philadelphia, PA, United States; Institute for Translational Medicine and Therapeutics, Perelman School of Medicine, University of Pennsylvania, Philadelphia, PA, United States; Department of Medicine, Perelman School of Medicine, University of Pennsylvania, Philadelphia, PA, United States; Department of Biostatistics, Epidemiology, and Informatics, Perelman School of Medicine, University of Pennsylvania, Philadelphia, PA, United States; Department of Translational Pharmacology, EWL School of Medicine, Bielefeld University, Bielefeld, Germany

## Abstract

**Objectives:** Non-steroidal anti-inflammatory drugs (NSAIDs) are recommended as first-line analgesics following third molar extraction, but some patients require supplemental opioids for pain management. The objective of this study was to identify demographic and clinical factors that predicted supplemental opioid use following third molar extraction in patients treated with an evidence-based analgesic regimen.

**Methods:** Healthy adults underwent surgical extraction of partial or full bony impacted mandibular third molar. When pain intensity was ≥4/10, participants were given ibuprofen 400 mg (N=59) or placebo (N=26) in a randomized, double-blind design. After 4h, all participants transitioned to open-label ibuprofen 400 mg + acetaminophen 500 mg, with oxycodone 5 mg available for breakthrough pain. Analgesic use was documented for the first week after extraction. Predictors of supplemental opioid use in addition to ibuprofen + acetaminophen were evaluated by logistic regression.

**Results:** Ibuprofen + acetaminophen provided adequate analgesia in most of the 85 participants, with 17 participants (20%) using supplemental oxycodone in the first week after extraction. Female sex (OR: 6.770; 95% CI: 1.657-35.57; p=0.013) and higher body mass index (BMI) (OR: 1.253; 95% CI: 1.052-1.525; p=0.016) were associated with increased odds of supplemental opioid use, while higher difficulty index (Pederson score) slightly decreased the odds of supplemental opioid use (OR: 0.852; 95% CI: 0.724-0.993; p=0.043). Adding pre-surgery neutrophil counts improved model fit, with higher neutrophil counts associated with lower odds of supplemental opioid use (OR: 0.435; 95% CI: 0.212-0.775; p=0.011).

**Conclusions:** Female sex, higher BMI, and pre-surgery neutrophil counts were predictors of supplemental opioid use in patients treated with an evidence-based analgesic regimen. Greater surgical difficulty of third molar extraction does not increase the likelihood of supplemental opioid use.

## Introduction

The extraction of third molar teeth is one of the most common reasons oral and maxillofacial surgeons (OMS) prescribe opioids. There is strong clinical trial evidence demonstrating the efficacy of non-steroidal anti-inflammatory drugs (NSAIDs) in relieving pain following surgical extraction of impacted third molars (Hersh et al. 2020; Miroshnychenko et al. 2023). Given the ongoing opioid epidemic, OMS should prescribe NSAIDs as first line analgesic therapy, unless contraindicated (Carrasco-Labra et al. 2024). However, some patients do not achieve sufficient pain relief with NSAIDs alone; in randomized clinical trials 20-30% patients required opioid rescue medication in addition to NSAID (Hersh et al. 2004; Hersh et al. 2000; Theken et al. 2019). OMS often prescribe small quantities of opioids “just-in-case” to ensure appropriate pain management in patients who will not respond adequately to NSAIDs (Moore and Hersh 2020).

One reason that is often used to justify prescription of an opioid for acute postoperative pain after third molar extraction is the potential for increased pain after a more difficult surgical procedure. Several studies indicate that greater surgical difficulty, defined either based on radiographic assessment or by length of surgery, is associated with greater pain intensity following third molar extraction (Ali et al. 2018; Benediktsdottir et al. 2004; Bortoluzzi et al. 2011; de Santana-Santos et al. 2013; Grossi et al. 2007; Lago-Mendez et al. 2007; Wang et al. 2017). Importantly, these studies have focused on identifying predictors of post-surgical pain intensity and have not investigated the impact on analgesic efficacy or supplemental opioid use.

Variability in the degree of pain relief by NSAIDs has long been recognized (Grosser et al. 2017), but to our knowledge there is little information about the mechanisms that might help to explain this finding. The innate immune response caused by tissue trauma initiates a plethora of molecular mechanisms that contribute to the pain sensation. Our previous work demonstrated that the analgesic response to ibuprofen following third molar extraction was associated with the degree of activation of the cyclooxygenase pathway, the molecular target of NSAIDs (Theken et al. 2019). Thus, clinical and demographic factors affecting these inflammatory processes, such as sex, age, leukocyte counts, and the extent of surgical tissue trauma, may contribute to variability in NSAID efficacy. We sought to identify clinical and demographic factors that predicted supplemental opioid use following third molar extraction in patients treated with an evidence-based NSAID-based analgesic regimen.

## Materials and Methods

The study consisted of two phases: a double-blind, placebo-controlled inpatient phase of the analgesic response to ibuprofen in the acute post-surgical period and an open-label outpatient phase in which all patients received the combination of ibuprofen and acetaminophen as their primary analgesic regimen. The primary endpoint was the use of opioid rescue medication, in addition to ibuprofen, in the first seven days following extraction. The study protocol was approved by the University of Pennsylvania Institutional Review Board (IRB#832417; ClinicalTrials.gov: NCT03893175), and all participants provided informed consent. The study was conducted in accordance with the Declaration of Helsinki.

Healthy volunteers (≥18 years of age) were recruited from patients referred to the Oral and Maxillofacial Surgery Service at the School of Dental Medicine and the Hospital of the University of Pennsylvania for extraction of one or more partially or fully bony impacted mandibular third molars between May 2019 and March 2022. Maxillary third molars were allowed in the surgical plan, if appropriate. Screening laboratory tests (complete metabolic panel and complete blood count with differential (CBC)) were performed in all participants approximately 1-2 weeks prior to surgery. Participants were excluded if they were obese (body mass index (BMI) >30 kg/m^2^); smoked or used recreational drugs; were pregnant or nursing a child; had a history of gastrointestinal ulcer, substance use disorder, diabetes, chronic pain, or inflammatory or autoimmune disease; were allergic or sensitive to NSAIDs or opioids; had contraindications to any of the study medications (ibuprofen, acetaminophen, or oxycodone); had an abnormal laboratory value that may be indicative of an underlying disease state; or had any serious medical condition that would make participation unsafe. Participants were asked to abstain from analgesics, including products containing NSAIDs, aspirin, and acetaminophen, high-dose vitamins, and nutritional supplements from enrollment until after surgery.

Third molar extraction was performed by one of four attending oral surgeons (N.P.; S.W.; R.A.H; and B.P.F.) using a standardized regimen of 2% lidocaine plus 1:100,000 epinephrine for local anesthesia with or without nitrous oxide/oxygen and/or midazolam + fentanyl (maximum 50 mcg) titrated to effect for sedation during the procedure and relatively rapid dissipation of the effect once surgery was complete. After surgery participants reported pain intensity every 15 minutes using the 0-10 Numeric Rating Scale (NRS-PI), where 0=no pain and 10=worst pain imaginable.

When the study participants reported a pain score ≥4/10 or indicated that their pain was no longer tolerable, they received a blinded dose of rapid-acting ibuprofen liquigel 400 mg or matching placebo by mouth, randomized in a ratio of 3:1. The blinded study medication was prepared by over encapsulation by the University of Pennsylvania Investigation Drug Service. Pain intensity assessments were continued every 15 minutes for 4 hours after study medication administration. Participants were encouraged to allow at least 60 minutes for the study medication to take effect before deciding if it had been ineffective, but rescue medication (oxycodone 5 mg) was allowed any time upon request.

Four hours after blinded study medication treatment, all participants received open-label ibuprofen 400 mg and acetaminophen 500 mg to be taken every 4 h around the clock for the first 2 days and then as needed for pain (Carrasco-Labra et al. 2024). Additionally, participants were given 8 oxycodone 5 mg tablets to be taken up to every 6 h as needed if they experienced insufficient pain relief on ibuprofen + acetaminophen. Participants received a diary to record their outpatient analgesic use and discharged home once medically stable. Participants returned the next day and on Day 7 after third molar extraction for verification of analgesic use and to return any unused study medication.

Study data were collected and managed using REDCap electronic data capture tools hosted at the University of Pennsylvania Perelman School of Medicine (Harris et al. 2019; Harris et al. 2009).

### Assessment of surgical difficulty

Surgical difficulty was evaluated by length of surgery from time of first incision to last suture, number of teeth extracted, trauma score, and Pederson scale. Trauma score, which incorporates both the number of teeth and degree of impaction, was calculated as previously described (maximum score=10) (Gordon et al. 1997). Each maxillary third molar was scored as 1 point. Mandibular third molars were scored as follows: partial bony impaction ≤50% bone coverage=1; partial bony impaction >50% bone coverage=2; full bony impaction, not horizontal=3, and full bony impaction, horizontal=4. Four attending oral surgeons (N.P.; S.W.; R.A.H; and B.P.F.) reviewed each patient’s panoramic radiograph to determine surgical difficulty for each molar based on the Pederson scale (Pedersen 1988). The median of the oral surgeons’ scores for each molar was used for subsequent analysis. The Pederson scores for each extracted tooth were summed to obtain an index of overall surgical difficulty for each patient.

### Statistical analysis

Demographic variables and measures of surgical difficulty are reported as mean ± standard deviation, median (interquartile range (IQR)) or percentages. Opioid rescue medication use during the inpatient phase between the ibuprofen and placebo groups was compared using Fisher’s exact test. Prediction modeling of the clinical and demographic factors associated with any opioid use in addition to ibuprofen (yes or no) throughout the study period was performed by logistic regression. Age and gender were included in all models to account for common covariates. Model 1 included demographic and clinical variables that are routinely available at the time of third molar extraction. Variables considered for inclusion in Model 1 were race, Hispanic ethnicity, BMI, summed Pederson score, length of surgery, trauma score, number of teeth extracted, use of IV sedation (midazolam + fentanyl) during the procedure, surgeon performing the procedure, and treatment (ibuprofen vs. placebo) during the double-blind inpatient phase. Model 2 included variables identified in Model 1 and additionally considered leukocyte counts from the screening CBC (white blood cells, neutrophils, monocytes, lymphocytes, eosinophils, and basophils). Variables were selected by stepwise regression including both forward selection and backward elimination to identify the model that optimized the Akaike information criterion (AIC). The final models were compared using the likelihood ratio test and assessed using area under the receiver operator characteristic (ROC) curve. Statistical analyses were performed in R (version 4.4.2).

## Results

Eighty-seven healthy adults were enrolled. Two patients did not report pain intensity ≥4/10 in the first 4 hours after third molar extraction and were excluded from further study procedures and analysis. During the double-blind, placebo-controlled inpatient phase, 59 patients received ibuprofen, and 26 patients received placebo. The demographic and clinical characteristics of the study cohort are shown in Table 1. Most third molar extractions were rated as moderately difficult according to the Pederson scale. The rate of post-surgical complications was low, with one patient experiencing a post-surgical infection and no cases of alveolar osteitis.

**Table 1:** Clinical and demographic characteristics of study cohort by supplemental opioid use in addition to ibuprofen.

|  | No Supplemental<br>Opioid Use (N=68) | Supplemental<br>Opioid Use (N=17) | Study Cohort<br>(N=85) |
| --- | --- | --- | --- |
| Men/Women (N (%)) | 34 (50%) / 34 (50%) | 5 (29.4%) / 12<br>(70.5%) | 39 (45.9 %) / 46<br>(54.1%) |
| Age (years (mean±SD)) | 24.2±3.8 | 23.9±2.9 | 24.2±3.6 |
| Race (N (%)) |  |  |  |
| American Indian/Alaska<br>Native | 1 (1.5%) | 0 (0%) | 1 (1.2%) |
| Asian | 27 (39.7%) | 6 (35.3%) | 33 (38.8%) |
| Black or African American | 13 (19.1%) | 5 (29.4 %) | 18 (21.2%) |
| White | 20 (29.4%) | 5 (29.4%) | 25 (29.4%) |
| Other | 7 (10.3%) | 1 (5.9%) | 8 (9.4%) |
| Hispanic ethnicity (N (%)) | 11 (16.2%) | 1 (5.9%) | 12 (14.1%) |
| Body Mass Index (kg/m<sup>2</sup><br>(mean±SD)) | 22.8±3.2 | 24.5±3.6 | 23.1±3.3 |
| Use of intravenous<br>sedation (N(%)) | 28 (41.2%) | 7 (41.2%) | 35 (41.2%) |
| Treatment during double-blind inpatient phase<br>Ibuprofen/Placebo (N (%)) | 46 (67.6%) / 22<br>(32.3%) | 13 (76.5%) / 4<br>(23.5%) | 59 (69.4%)/ 26<br>(30.6%) |
| Length of surgery (minutes<br>(mean±SD)) | 28.6±16.2 | 28.6±14.0 | 28.6±15.7 |
| Number of teeth extracted<br>(median (IQR)) | 4 (2.25, 4) | 4 (2.5, 4) | 4 (2.5,4) |
| Trauma score (median<br>(IQR)) | 6.5 (4, 8) | 6 (4, 8.5) | 6 (4,8) |
| Pederson Score (median<br>(IQR; n)) |  |  |  |
| Tooth #1 | 5.0 (4.0, 6.0; n=59) | 5.5 (4.0, 6.0; n=14) | 5.0 (4.0, 6.0;<br>n=73) |
| Tooth #16 | 5.0 (4.0, 6.0; n=64) | 6.0 (5.125, 6.25;<br>n=14) | 5.5 (4.0, 6.0;<br>n=78) |
| Tooth #17 | 6.0 (5.5, 7.0; n=68) | 6.0 (5.0, 6.75; n=17) | 6.0 (5.5, 6.875;<br>n=85) |
| Tooth #32 | 6.0 (5.0, 7.0; n=65) | 5.75 (5.0, 6.5; n=16) | 6.0 (5.0, 7.0;<br>n=81) |
| Summed Pederson Score | 21.5 (19.5, 23.5) | 21.5 (17.0, 28.5) | 21.5 (19.5, 24.0) |
| White Blood Cell Counts ( x<br>10<sup>3</sup>/μl (mean±SD)) | 6.21±1.72 | 5.00±1.48 | 5.97±1.74 |
| Neutrophil Counts ( x 10<sup>3</sup>/μl<br>(mean±SD)) | 3.74±1.40 | 2.62±1.38 | 3.52±1.46 |
| Monocyte Counts ( x 10<sup>3</sup>/μl<br>(mean±SD)) | 0.44±0.14 | 0.36±0.13 | 0.43±0.14 |
| Lymphocyte Counts ( x<br>10<sup>3</sup>/μl (mean±SD)) | 1.89±0.45 | 1.87±0.37 | 1.88±0.43 |
| Eosinophil Counts ( x 10<sup>3</sup>/μl (mean±SD)) | 0.11±0.11 | 0.11±0.09 | 0.11±0.10 |
| Basophil Counts ( x 10<sup>3</sup>/μl (mean±SD)) | 0.02±0.04 | 0.01±0.02 | 0.02±0.04 |

Ibuprofen was significantly more effective than placebo in reducing pain in the acute post-surgical period, with 3 patients in the ibuprofen-treated group requesting opioid rescue medication (5.1%) during the 4-hour inpatient observation period compared to 21 patients who received placebo (80.8%, p<0.05). The combination of ibuprofen 400 mg + acetaminophen 500 mg provided adequate pain control in most patients during the first week following extraction, with 16 of the 85 patients (18.8%) using any oxycodone during the open-label outpatient phase. Of the 17 patients in total who used supplemental oxycodone in addition to ibuprofen, the median number of pills consumed was 3 (2, 7.5).

Modeling results and ROC curves are shown in Table 2 and Figure 2, respectively. Model 1 retained age, gender, BMI, and summed Pederson index as predictors of opioid use in addition to ibuprofen, with an area under the ROC curve of 0.738 (0.588-0.889). Addition of pre-surgery neutrophil counts (Model 2) significantly improved model fit (p=0.003), with an area under the ROC curve of 0.806 (0.665-0.947). Using a predicted probability cut-off of 0.3, Model 1 had a sensitivity of 52.9% and a specificity of 83.6% for predicting supplemental opioid use, with a positive predictive value of 45.0% and a negative predictive value of 87.5%. Model 2 had a sensitivity of 64.7% and a specificity of 88.1% for predicting supplemental opioid use, with a positive predictive value of 57.9% and a negative predictive value of 90.8%.

**Table 2:** Logistic regression results.

|  | Odds ratio (95% CI) | p-value |
| --- | --- | --- |
| Model 1 |  |  |
| Age (years) | 0.933 (0.765-1.106) | 0.451 |
| Female Gender | 6.770 (1.657-35.57) | 0.013 |
| BMI | 1.253 (1.052-1.525) | 0.016 |
| Summed Pederson Score | 0.852 (0.724-0.993) | 0.043 |
| Model 2 |  |  |
| Age (years) | 0.924 (0.734-1.136) | 0.469 |
| Female Gender | 6.522 (1.536-36.16) | 0.018 |
| BMI | 1.273 (1.046-1.595) | 0.023 |
| Summed Pederson Score | 0.910 (0.763-1.076) | 0.275 |
| Neutrophil Counts | 0.435 (0.212-0.775) | 0.011 |

**Figure 1:**
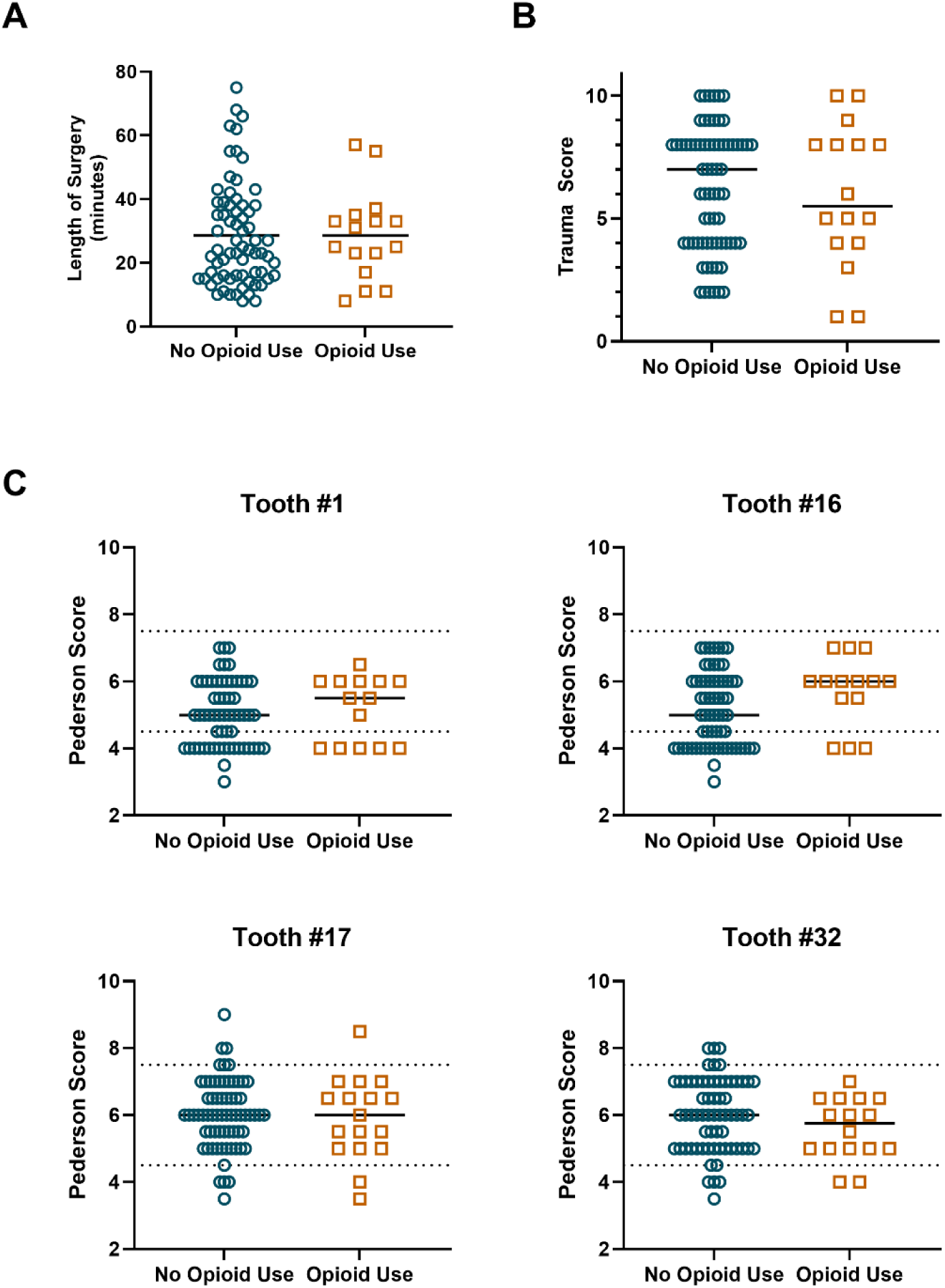
Comparison of (A) length of surgery, (B) trauma score, and (C) Pederson index for each extracted third molar between patients who achieved adequate pain relief with ibuprofen (No Opioid Use) and patients who required oxycodone in addition to ibuprofen (Opioid Use). The dashed lines indicate the thresholds for minimally difficult, moderately difficult, and very difficult.

**Figure 2:**
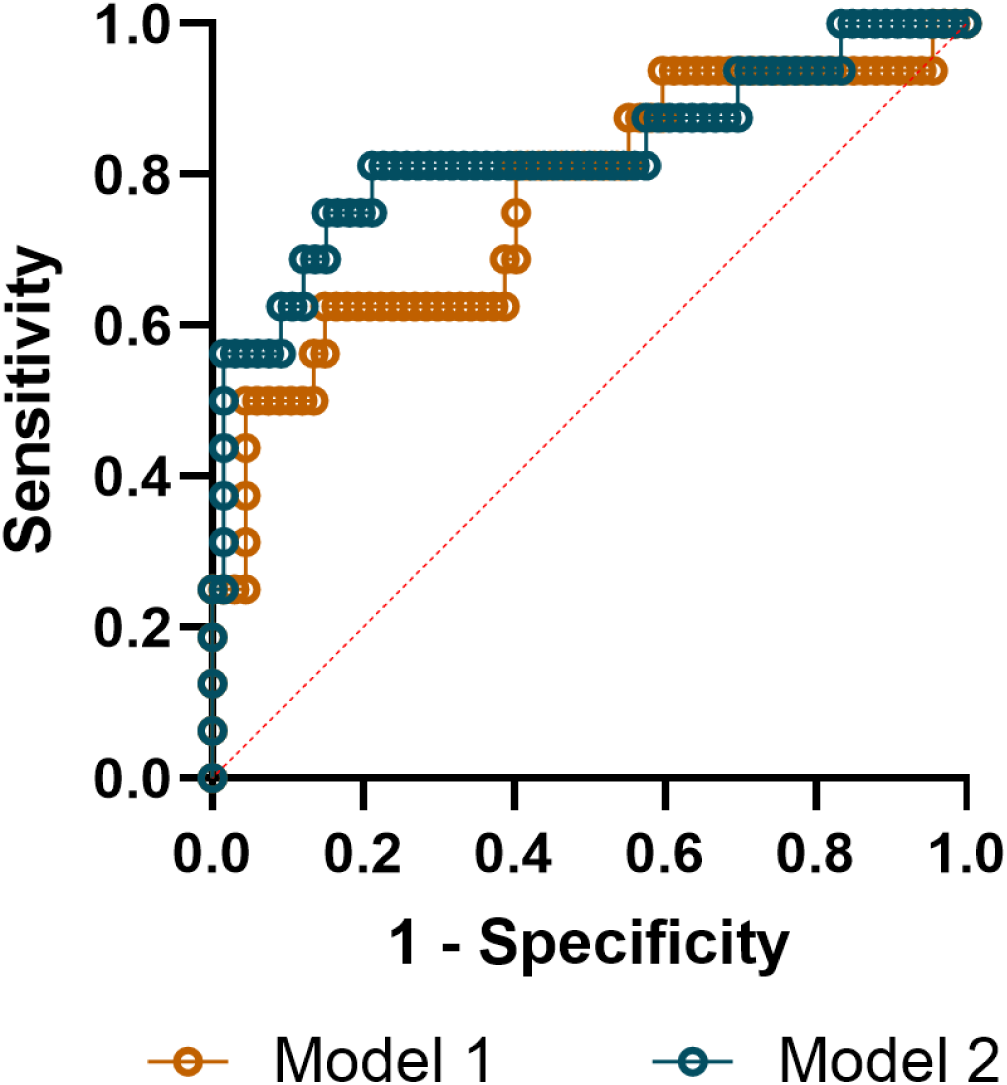
Receiver operator characteristic curves of Model 1 (age, gender, BMI, and summed Pederson index) and Model 2 (age, gender, BMI, summed Pederson index, and pre-surgery neutrophil counts).

## Discussion

We observed that the combination of ibuprofen and acetaminophen was very effective for management of post-operative pain following third molar extraction, as would be anticipated from prior randomized clinical trials (Feldman et al. 2025; Miroshnychenko et al. 2023). However, 20% of patients in our cohort used supplemental oxycodone in the first week after extraction, requiring a median of 3 pills. Similarly, a previously published prospective study in adolescents and young adults indicated that opioid use was minimal following extraction of four third molars, with 7% of patients reporting use of oxycodone 5 mg and consuming a mean of 3.3 pills per patient (Resnick et al. 2019). These findings support recommendations that empiric opioid prescribing is not necessary for all patients undergoing third molar extraction and, when needed, the number of opioid pills prescribed should be limited (Carrasco-Labra et al. 2024).

While mandatory use of prescription drug monitoring programs and electronic health record defaults have decreased the quantities of opioids prescribed by dentists (Bachhuber et al. 2023; Wang et al. 2021), these strategies do not address opioid prescriptions written “just-in-case” (Moore and Hersh 2020). To our knowledge, this is the first study to show that clinical and demographic factors predict supplemental opioid use in patients receiving an evidence-based analgesic regimen after third molar extraction. A model incorporating age, gender, BMI, and Pederson index demonstrated good predictive performance, which improved further with the addition of pre-surgery neutrophil counts. These findings provide a foundation for developing precision medicine approaches to managing post-surgical dental pain.

We found that pre-surgery neutrophil counts predicted insufficient pain relief with ibuprofen + acetaminophen, with higher neutrophil counts associated with lower odds of supplemental opioid use. Of note, individuals with active infection or chronic inflammatory conditions were excluded from our study, and most participants had pre-surgery neutrophil counts within the reference range for healthy adults. Neutrophils are the first circulating inflammatory cells recruited to the site of injury and promote acute pain, in part via production of PGE_2_ (Cunha et al. 2008). Our prior work demonstrated that participants who used opioid in addition to ibuprofen exhibited lower production of PGE_2_ *in vivo* compared to participants who achieved adequate pain relief with ibuprofen alone (Theken et al. 2019). Our current findings support these observations, suggesting that neutrophils are a key contributor to cyclooxygenase pathway activation following third molar extraction and that a robust acute inflammatory response to surgical trauma determines analgesic efficacy of NSAIDs. A recent study in a human model of incisional pain reported that enhanced neutrophil infiltration was associated with greater hyperalgesia after incision (Segelcke et al. 2023). However, other studies suggest that depletion of neutrophils can promote persistent pain (Parisien et al. 2022). Thus, additional studies are warranted to define the role of neutrophils in mediating post-surgical pain and analgesic response to NSAIDs.

We also found that female gender increased the odds of supplemental opioid use. It is well-recognized that sex impacts the immune response (Klein and Flanagan 2016), which may contribute to sex differences in inflammatory pain. Prior studies reported an association between gender and absolute post-surgical pain intensity, with most indicating that women report higher pain scores (Benediktsdottir et al. 2004; de Santana-Santos et al. 2013; Phillips et al. 2003). Other studies have reported greater pain intensity in men (Capuzzi et al. 1994) or no association between gender and post-surgical pain (Ali et al. 2018; Bortoluzzi et al. 2011). These conflicting results could be due in part to differences in the timing of pain assessments or the scales used, as some used a 0-10 NRS while others used a 4-point categorical scale (none, mild, moderate, severe). However, few studies have investigated gender differences in analgesic response to NSAIDs. One study in a model of experimental pain induced by electrical stimulation reported a greater analgesic response to ibuprofen in men compared to women (Walker and Carmody 1998). In contrast, a meta-analysis of studies of third molar extraction pain submitted to the Food and Drug Administration (FDA) found no gender differences in mean pain intensity scores and mean pain relief scores with ibuprofen (400 mg) in the first 6 hours after drug administration. They noted some heterogeneity among the included studies, with 5 reporting no gender differences, 1 study reporting lower pain intensity after medication in women, and 1 reporting lower pain intensity after medication in men (Averbuch and Katzper 2000). Although similar in design and enrollment criteria, these studies each included a relatively small number of patients (approximately N=45), evaluated analgesic response only in the acute post-surgical period (first 6 hours), and were not designed to investigate gender differences in pain relief. Additional studies are necessary to clarify the effects of gender on analgesic response to NSAIDs in post-surgical pain.

Higher BMI was also associated with an increased odds of opioid use in addition to ibuprofen + acetaminophen. This is consistent with prior studies demonstrating that BMI was associated with both post-operative pain intensity and opioid requirements in a variety of surgical models (Abrecht et al. 2019; Yang et al. 2019). However, none of these studies have specifically evaluated the impact of overweight or obesity on analgesic response to NSAIDs. Obesity can affect drug disposition, and lower ibuprofen peak plasma levels have been reported in obese individuals (BMI: 38.6±3.3 kg/m^2^) compared to control individuals (BMI: 20.7±0.5 kg/m^2^) (Abernethy and Greenblatt 1985). However, obese subjects were excluded from our study cohort, and we did not observe any relationship between BMI and degree of COX-1 or COX-2 inhibition following ibuprofen treatment in our cohort (data not shown), suggesting that the effect of BMI on supplemental opioid use is not mediated by differences in pharmacokinetics.

Difficulty and length of the surgical procedure are often taken into consideration when deciding whether to prescribe opioids. Prior studies reported that greater surgical difficulty, as assessed by tooth position and morphology, a 4-class rating scale (I, extraction requiring forceps only; II, extraction requiring osteotomy; III, extraction requiring osteotomy and coronal section; IV, complex extraction (root section)), and/or length of surgery, was associated with greater pain intensity following third molar extraction (Ali et al. 2018; Bortoluzzi et al. 2011; de Santana-Santos et al. 2013; Grossi et al. 2007; Lago-Mendez et al. 2007; Wang et al. 2017). However, none of these studies investigated whether surgical difficulty impacted the degree of pain relief patients experienced when treated with evidence-based analgesia. Our results suggest that greater surgical difficulty is not a robust indicator of which patients will require a supplemental opioid prescription for optimal pain relief after third molar extraction. Interestingly, we observed that greater Pederson index was associated with minimally lower odds of supplemental opioid use. Some studies have questioned whether the Pederson index is a reliable measure of surgical difficulty (Bali et al. 2013). Therefore, we also considered other metrics, including number of teeth, a trauma score that incorporates the number of teeth and degree of impaction, and length of surgery. However, none of these were associated with supplemental opioid use in the first week following extraction.

While our predictive model exhibited excellent predictive performance, it correctly classified only 64.7% of patients who used opioids. These results suggest that clinical factors alone cannot completely predict the need for supplemental opioids following third molar extraction. Numerous biological, psychological, and social mechanisms can contribute to and modulate pain perception (Vardeh et al. 2016). Thus, it is not surprising that there is substantial inter-individual variability in both pain intensity and analgesic response following surgery. Further studies designed to parse the diverse mechanisms underlying variability in analgesic response are warranted.

Our study has several limitations. Although we recruited a diverse cohort, our results were obtained from patients at a single clinical center and may not be generalizable to other sites. Our study cohort was also limited in size, which precludes a comprehensive analysis of all the factors that might contribute to heterogeneity in the analgesic response to ibuprofen. We enrolled only patients with ≥50% partial bony impaction of at least one mandibular third molar, so our results may differ from what would be observed in patients undergoing nonsurgical extractions or with lesser degrees of impaction. Finally, the use of corticosteroids or long-acting local anesthetics was not permitted in our study protocol. Thus, our results may not reflect what would be seen in clinical practice where these interventions are commonly used.

## Conclusions

In conclusion, while our study revealed pre-surgery neutrophil counts, gender, and BMI as predictors of NSAID analgesic response, indices of greater surgical difficulty were not a robust predictor of whether a patient will require a supplemental opioid for pain relief when receiving a standard-of-care NSAID-based analgesic regimen following third molar extraction. Additional studies are necessary to elucidate the mechanisms that contribute to variability in analgesic response to NSAIDs and need for opioids following third molar extraction.

## Data Availability

All data produced in the present study are available upon reasonable request to the authors

## Acknowledgements

This work was supported by funds from the Penn Center for Precision Medicine, an Oral and Maxillofacial Surgery Foundation Clinical Research Grant, and funding from the National Institute of Oral and Craniofacial Research (R01DE033405) and the National Center for Advancing Translational Sciences of the National Institutes of Health (UL1TR001878). The content is solely the responsibility of the authors and does not necessarily represent the official views of the National Institutes of Health. The funders had no role in the study design, in the collection, analysis and interpretation of data; in the writing of the manuscript; or in the decision to submit the manuscript for publication.

## Author Contributions

Neeraj Panchal: Contributed to patient recruitment, performed third molar extractions, contributed to data acquisition and interpretation, drafted, and critically revised the manuscript Steven Wang: Contributed to patient recruitment, performed third molar extractions, contributed to data acquisition and critically revised the manuscript

Rania A. Habib: Contributed to patient recruitment, performed third molar extractions, contributed to data acquisition and critically revised the manuscript

Brian P. Ford: Contributed to patient recruitment, performed third molar extractions, contributed to data acquisition and critically revised the manuscript.

Truongan Pham: Contributed to data acquisition and critically revised the manuscript.

Perla M. Cherfane: Contributed to data acquisition and critically revised the manuscript.

Gaurav Gupta: Contributed to data acquisition and critically revised the manuscript.

Sydney A. Juska: Contributed to data acquisition and critically revised the manuscript.

Stacey A. Secreto: Contributed to patient recruitment, data acquisition, and critically revised the manuscript.

Gregory R. Grant: Contributed to conception, design, data acquisition and interpretation, and critically revised the manuscript

Carsten Skarke: Contributed to conception, design, data acquisition and interpretation, and critically revised the manuscript.

Elliot V. Hersh: Contributed to conception, design, data interpretation, and critically revised the manuscript.

John T. Farrar: Contributed to conception, design, data interpretation, and critically revised the manuscript.

Tilo Grosser: Contributed to conception, design, data acquisition and interpretation, and critically revised the manuscript.

Katherine N. Theken: Contributed to conception, design, data acquisition and interpretation, performed statistical analyses, drafted, and critically revised the manuscript.

All authors gave their final approval and agree to be accountable for all aspects of the work.

## Disclosures

The authors have no conflicts of interest to disclose.

## Notes

### Competing Interest Statement

The authors have declared no competing interest.

### Clinical Trial

NCT03893175

### Author Declarations

The study protocol was approved by the University of Pennsylvania Institutional Review Board (IRB#832417), and all participants provided informed consent.

